# Development of Longitudinal, Linked Maternal-Infant Cohorts using the Epic Cosmos Electronic Health Record Dataset

**DOI:** 10.64898/2026.06.02.26354757

**Authors:** Stephanie A. Leonard, Kevin Dysart, Alison Callahan, Sara Siadat, Jiaqi Zhang, Sara C. Handley, Krista F. Huybrechts, Irogue Igbinosa, Brian T. Bateman

## Abstract

**Background:** Epic Cosmos is a relatively new centralized dataset of electronic health records, largely in the U.S., with high potential utility in perinatal epidemiologic research.

**Objectives:** The study objectives were to develop replicable steps to create longitudinal, linked maternal-infant cohorts in Cosmos, assess completeness of key variables, evaluate potential selection bias with restrictions for longitudinal healthcare encounters, and provide an example epidemiologic analysis.

**Methods:** We created maternal-infant cohorts by starting with infants born during 2023-2024 and joining with additional data tables as needed. We selected and created variables for perinatal characteristics, common comorbidities, and routinely measured vital signs and laboratory values, and assessed variable completeness. We sequentially restricted the birth cohort for maternal-infant linkage and longitudinal healthcare from first-trimester prenatal care encounter through infant follow-up care within 12 weeks post-discharge from birth hospitalization. Finally, we conducted an example analysis of the association between high systolic blood pressure in the first trimester (≥140 mm Hg) and later onset of preeclampsia among those with chronic hypertension.

**Results:** The total linked birth cohort included 2,929,600 pregnancies. Completeness was >90% for most variables assessed but was 76% for racial and ethnic group and 77% for body mass index at delivery. Characteristics of the cohort were similar to those reported for the entire United States birth population based on birth certificate data, including similar regional and racial-ethnic composition. Longitudinal cohort restriction requiring linked records from first trimester prenatal care through infant follow-up care reduced the cohort size to 586,841 pregnancies. However, restriction had minimal effects on cohort characteristics. In the example analysis, high systolic blood pressure was associated with increased risk of preeclampsia among those with chronic hypertension (aRR: 1.26; 95% CI: 1.23, 1.30).

**Conclusions:** This study provides a rigorous and reproducible approach to creating longitudinal, linked maternal-infant cohorts in Epic Cosmos. The analytical findings suggest high data quality and representativeness of the U.S., while also drawing attention to dataset limitations that perinatal epidemiologic researchers should be aware of, notably attrition with longitudinal restrictions.

**SYNOPSIS:** • **Study question:** The study sought to develop replicable steps for researchers to create longitudinal, linked maternal-infant cohorts using the Epic Cosmos electronic health record (EHR) dataset, assess completeness of key variables, evaluate potential selection bias with sequential restrictions for longitudinal healthcare encounters, and provide an example analysis.

• **What’s already known:** Epic Cosmos is a relatively new dataset with high potential for perinatal epidemiologic research because of its large size and granular information from structured EHR data.

• **What this study adds:** This study provides a rigorous and reproducible approach to creating longitudinal, linked maternal-infant cohorts in Epic Cosmos and found overall high completeness of key variables, and high representativeness of the United States. Cohort characteristics did not meaningfully change with longitudinal cohort selection.

## BACKGROUND

Electronic health record (EHR) systems have been widely adopted in clinical practice to document patients’ conditions, tests, and treatments^1^ and contain granular, longitudinal health information collected in real-world clinical settings. These data are potentially a rich source for epidemiologic studies, but concerns about data quality and representativeness have limited their use.^2–4^ Data quality concerns include missing measurements and encounters. Representativeness concerns stem from the fact that EHR based studies typically rely on data from a small number of healthcare organizations. Combining EHR data from many institutions for research purposes has historically been a major challenge. Further, perinatal epidemiologic research frequently requires linkage of maternal and infant records, which is challenging in many EHR data sources.

Epic Cosmos is a relatively new dataset that centralizes EHR data from hundreds of healthcare organizations.^5^ Epic is the leading EHR software vendor in the United States, and healthcare organizations that use Epic Systems’ EHR can now opt to contribute to Cosmos and access the Cosmos dataset. If a healthcare organization participates in Cosmos, investigators affiliated with the organization can access the dataset after completing required trainings, with investigator costs varying depending on the level of training and project funding. In Cosmos, information is pulled from all structured EHR fields, with high granularity of patient health information, including laboratory measurements, vital signs, and medications prescribed or administered. Organizations participating in Cosmos range from academic to community health systems, are located throughout the United States and select locations in Saudi Arabia, Lebanon, and Canada (currently), and continue to grow in number. The Cosmos dataset currently contains data from approximately 300 million patients seen at over 2,000 hospitals and 47,000 clinics.^5^

Interest in and use of Epic Cosmos for perinatal epidemiologic research are growing rapidly, creating a critical need to assess data quality and representativeness of maternal-infant cohorts in Cosmos.^6–8^ The goal of this study was to provide detailed, replicable steps to create longitudinal, linked maternal-infant cohorts in Cosmos, assess completeness of key variables, and evaluate potential selection bias via restrictions for longitudinal healthcare encounters. In addition, we sought to provide perinatal epidemiologic researchers with an applied example, by using the Cosmos dataset to estimate the association between systolic blood pressure (SBP) in the first trimester of pregnancy and later onset of preeclampsia among those with chronic hypertension. Prior studies in selective study samples have found that individuals with higher SBP in the first trimester were more likely to develop preeclampsia.^9–11^ Screening for high SBP and modifying antihypertensive treatment early in pregnancy could be potential strategies to reduce the risk of preeclampsia, which affects approximately 30% of women with chronic hypertension and poses considerable risks to the pregnant woman and infant.^12–14^

## METHODS

### Cohort Creation

Steps taken for cohort creation are described here and in greater detail in Appendix 1. We first selected live births recorded in the Cosmos birth records from January 1, 2023 to December 31, 2024. The data table of births is populated for a patient when an EHR is created for an infant and documentation is made in the delivery summary. New EHRs are typically only created for liveborn infants and the existence of a delivery summary indicates that the delivery occurred in the same healthcare organization that is contributing the data to Epic Cosmos. Therefore, pregnancies resulting in non-livebirths, including stillbirths, spontaneous and therapeutic abortions, and ectopic pregnancies, and those delivered at an organization that does not participate in Cosmos were not included. Such pregnancies would need to be identified using a different approach based on other data tables, including but not limited to the pregnancy data table. We selected 2023 to 2024 to have complete years of data and infant follow-up through 12 weeks post-discharge from birth hospitalization at the time of analysis, but the methods described here can be implemented for all available years in Cosmos when accounting for the necessary follow-up time in a given study. Birth data available in Cosmos are considered reliable as early as 2017 or when an organization started using Epic, whichever occurred later. Data are updated in Cosmos approximately every two weeks and we conducted analyses using data updated through 9/2/2026.

We then linked the birth records to pregnancy records based on a maternal patient identifier. Pregnancy records are populated when a patient receives healthcare during their pregnancy. A birth record is unique to each liveborn infant; for example, if an obstetric patient gives birth to liveborn twins, there will be two birth records and one pregnancy record. To link to birth encounters during the same birth years (2023-2024) we joined the existing dataset with records for each encounter that a patient had with the healthcare organization that was documented in an EHR or from billing data. We then removed unavailable birth encounters (i.e., missing). Birth records can be populated retrospectively by a clinician at a later date and not be linked to a birth encounter. Next, we identified twins and higher-order multiple births and, if multiple birth dates, assigned the earlier date as the delivery date of the pregnancy. In some instances, a stillborn infant will have an EHR created, which will be captured as a live birth record. We therefore excluded any pregnancies with an ICD code indicative of stillbirth.

### Study Variables

We selected and created variables that are frequently used in perinatal epidemiologic research and surveillance. All variables are detailed in **Appendix 2**, including time windows used for diagnoses, procedures, or measurements. We additionally created variables that are important in obstetric or neonatal health but not typically available or reliable in administrative claims or vital records data. For example, parity, body mass index (BMI), infant birthweight, and Apgar score are not well-captured in administrative claims data; hypertensive disorders, diabetes, and depression are not well-captured in vital records data; and blood pressure measurements and hemoglobin values are not well-captured in either administrative claims or vital records data. For gestational age, if one pregnancy had multiple gestational ages, we assigned the lowest. If no gestational age was available, we calculated it as the delivery date from the birth record minus the estimated start date from the pregnancy record. If that was missing, we set gestational age to missing. We calculated the last menstrual period as the delivery date minus the gestational age.

### Longitudinal, Linked Maternal-Infant Cohorts

We then placed restrictions to identify maternal-infant dyads with continuity of healthcare capture in Epic beyond the birth hospitalization. Some patients give birth at hospitals that use Epic Systems for EHRs and participate in Cosmos, but receive their prenatal care in clinics that do not use Epic or do not participate in Cosmos such that these encounters are not captured in the Cosmos database. Excluding such patients is necessary for research questions with exposures, outcomes, or covariates that occur before or after the birth hospitalization, but reduce cohort size and therefore statistical power and may also induce selection bias. To examine this phenomenon, we started with the birth cohort and then added sequential restrictions based on encounter type, department of encounter, and timing of encounter relative to the start of pregnancy or date of delivery. First, we created a cohort with likely continuous healthcare documented in Epic throughout pregnancy and birth by requiring at least one prenatal care encounter in the first trimester (before gestational week 14) and at least one prenatal care encounter in the second trimester (from gestational week 14 to week 27). Second, we extended the restriction to require at least one postpartum care encounter within 12 weeks of delivery. Third, we added a requirement of at least one infant follow-up care encounter within 12 weeks of discharge from the birth hospitalization. These steps are further detailed in Appendix 3.

### Statistical Analysis

We first calculated the frequencies and percentages of completeness for key perinatal variables. Next, we calculated the frequencies and percentages of maternal and neonatal characteristics in each of the four cohorts with increasingly restrictive longitudinal selection: (1) linked maternal-infant birth encounters, (2) birth and prenatal care encounters, (3) birth, prenatal, and postpartum care encounters, (4) birth, prenatal, postpartum, and infant follow-up care encounters. Finally, we conducted an example analysis, replicating the known association between high SBP in the first trimester of pregnancy and later onset of preeclampsia among pregnant individuals with chronic hypertension. For this analysis, we selected individuals with at least one prenatal care encounter in the first trimester from the total birth cohort. We then restricted to individuals with a diagnosis of chronic hypertension between three months prior to pregnancy and 20 weeks’ gestation and a SBP recorded during a first trimester prenatal care encounter. We defined high SBP as ≥140 mm Hg following hypertension guidelines of the American College of Obstetricians and Gynecologists.^15^ We estimated risk ratios (RR) with 95% confidence intervals (CI) using multivariable modified Poisson regression models with robust standard errors. We used multiple imputation by chained equations (50 imputations) to account for missing values of the covariates: maternal age (0.04%), insurance status (6.4%), and race-ethnicity (19.4%). Analysis was conducted in R version 4.6.1.

## RESULTS

The total cohort included 3,009,596 pregnancies to 2,944,291 mothers and 3,899,013 infants (**Figure 1**). After restricting to linked maternal-infant records, there was a 3% reduction in the number of pregnancies and mothers and a 24% reduction in the number of infants. Maternal-infant linkage in the birth cohort requires that the infant was delivered at a Cosmos-participating healthcare organization, has a mother documented in their family history, and the delivery episode was recorded in the mother’s EHR. Therefore, an infant who received healthcare at a Cosmos-participating hospital or clinic but was born at a non-participating hospital could not be linked to a maternal record. Restricting the cohort to those with prenatal care encounters documented during the first and second trimesters, reduced the cohort size by an additional 62%. The excluded patients would be those who received prenatal care at a healthcare organization that does not participate in Epic Cosmos and those who received delayed or no prenatal care. Restricting the cohort to those with a postpartum visit, reduced the cohort size by 16%. Finally, requiring that there be a linked infant follow-up visit, reduced the cohort size by 40%. The cohort sizes in this most restrictive linked, longitudinal cohort were 577,688 pregnancies, 568,411 mothers, and 586,841 infants during the two-year study period. The cohort sizes differ for each group because a small proportion (∼1.5%) of mothers had more than one pregnancy during the two years and a small proportion (∼1.5%) of pregnancies were twins or higher-order multiples.

**Figure 1.**
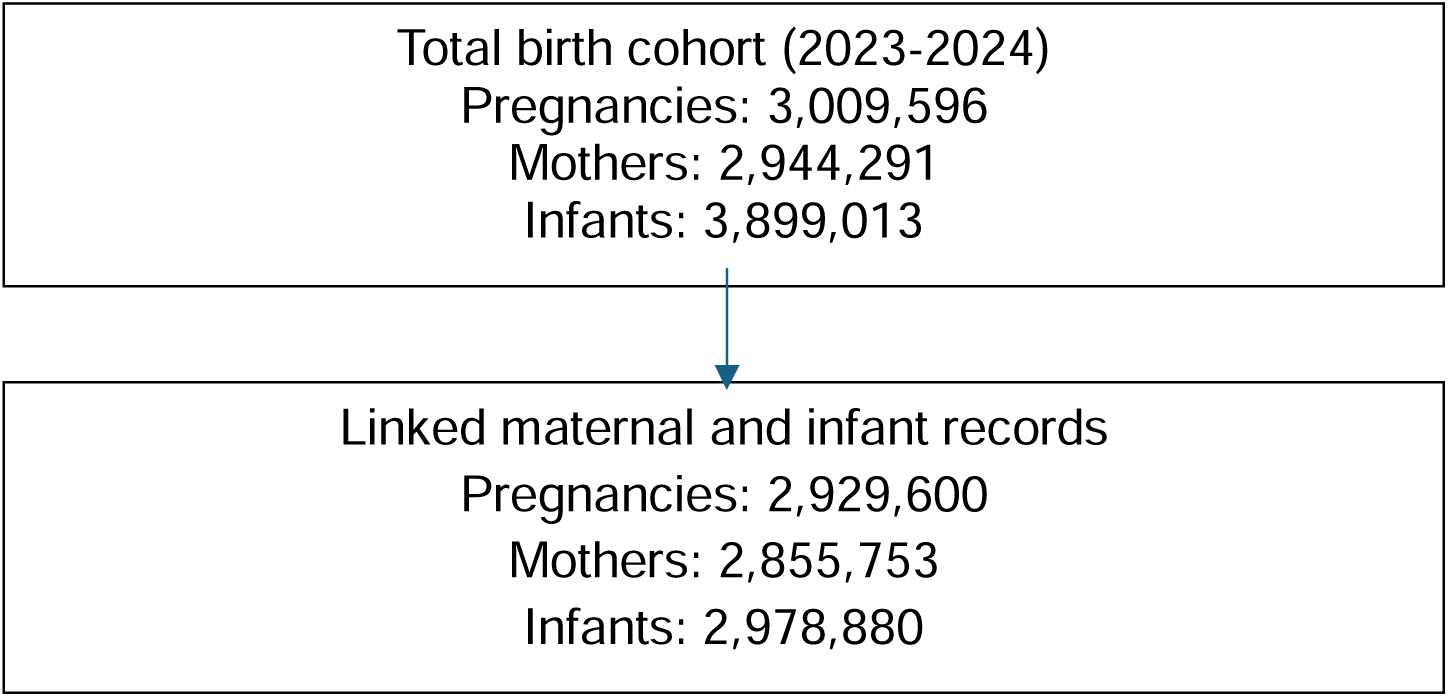
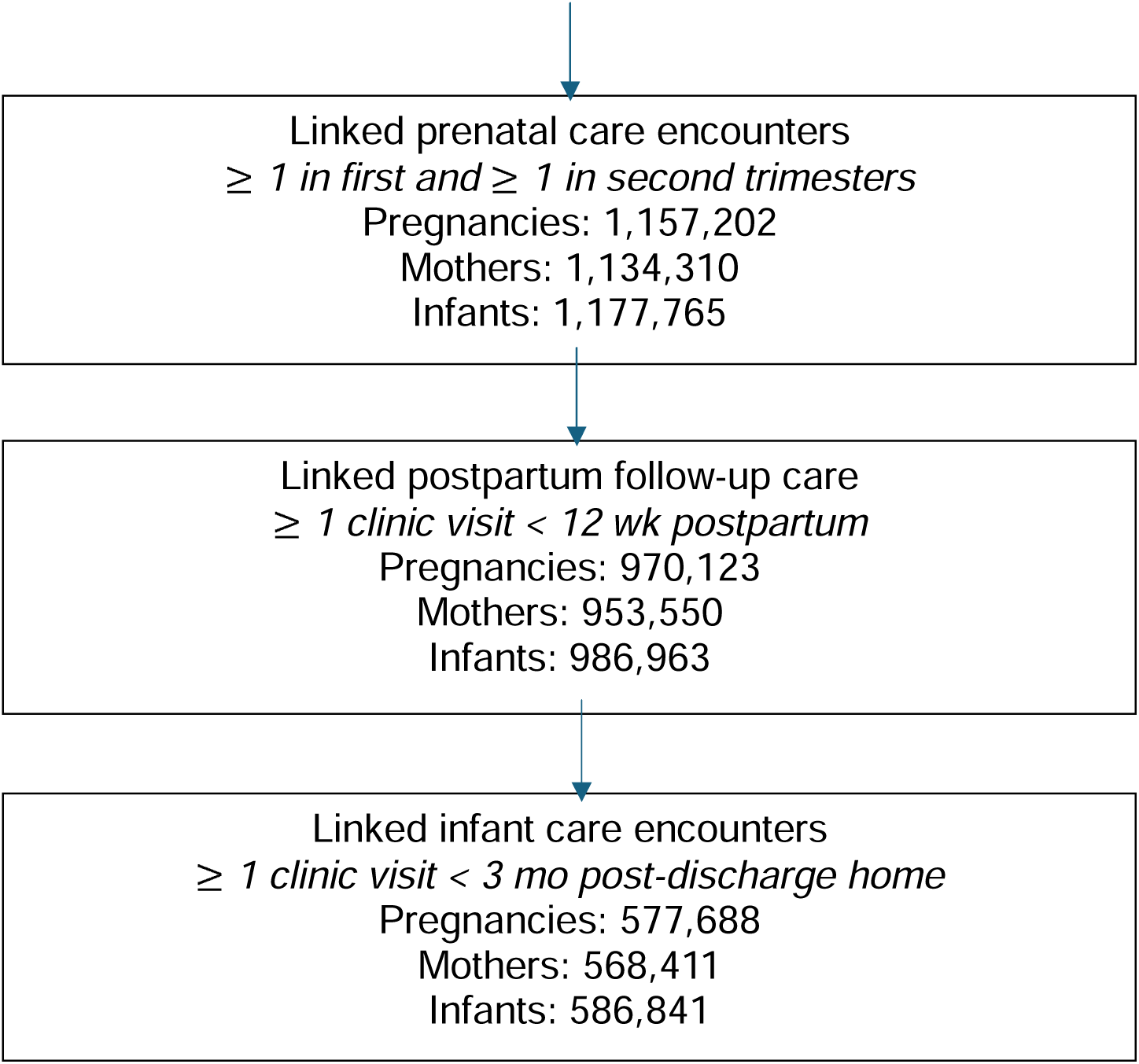
Linked, longitudinal birth cohort selection

Data completeness varied among key perinatal variables (**Table 1**). Completeness was >97% for age, region of residence, rurality, parity, multiple gestation, gestational age, mode of birth, birthweight, and 5-minute Apgar score. It was lower (89-92%) for hemoglobin and blood pressure measurements during delivery hospitalization and for payer status. Completeness was lowest for racial-ethnic group (76%) and BMI at delivery (77%).

**Table 1.** Completeness of characteristics (N = 2,978,880 infant records in linked maternal-infant cohort)

| Characteristics | N (%) |
| --- | --- |
| Maternal age at delivery | 2,978,879 (99.9) |
| Racial and ethnic group | 2,277,875 (76.5) |
| Payer for delivery encounter | 2,692,502 (90.4) |
| Region | 2,948,548 (99.0) |
| Rurality | 2,895,328 (97.2) |
| Delivery BMI | 2,284,544 (76.7) |
| Parity | 2,978,880 (100.0) |
| Hemoglobin at delivery admission | 2,753,446 (92.4) |
| Blood pressure at delivery admission | 2,662,895 (89.4) |
| Multiple gestation | 2,662,895 (89.4) |
| Gestational age at birth | 2,978,880 (100.0) |
| Mode of birth | 2,973,460 (99.8) |
| Birthweight | 2,949,881 (99.0) |
| Apgar score at 5 minutes | 2,975,605 (99.9) |

Maternal health characteristics were similar across the four cohorts with increasingly restrictive longitudinal records (**Table 2**). The cohorts were diverse with respect to racial-ethnic group, payer status, and region of residence in the United States. The mean maternal age was 30 years old and most individuals lived in metropolitan areas, were multiparous, had a BMI ≥30 kg/m^2^ at delivery, and gave birth at a community hospital. In the total linked birth cohort, the prevalence of common morbidities was 11% for preeclampsia, 9% for gestational diabetes, 12% for depression, and 25% for anemia. At admission for delivery, mean hemoglobin was 11.8 g/dL (standard deviation (SD): 1.3) and mean blood pressure was 126/77 mm Hg (SD: 14.7 for systolic BP and 11.1 for diastolic BP).

**Table 2.**
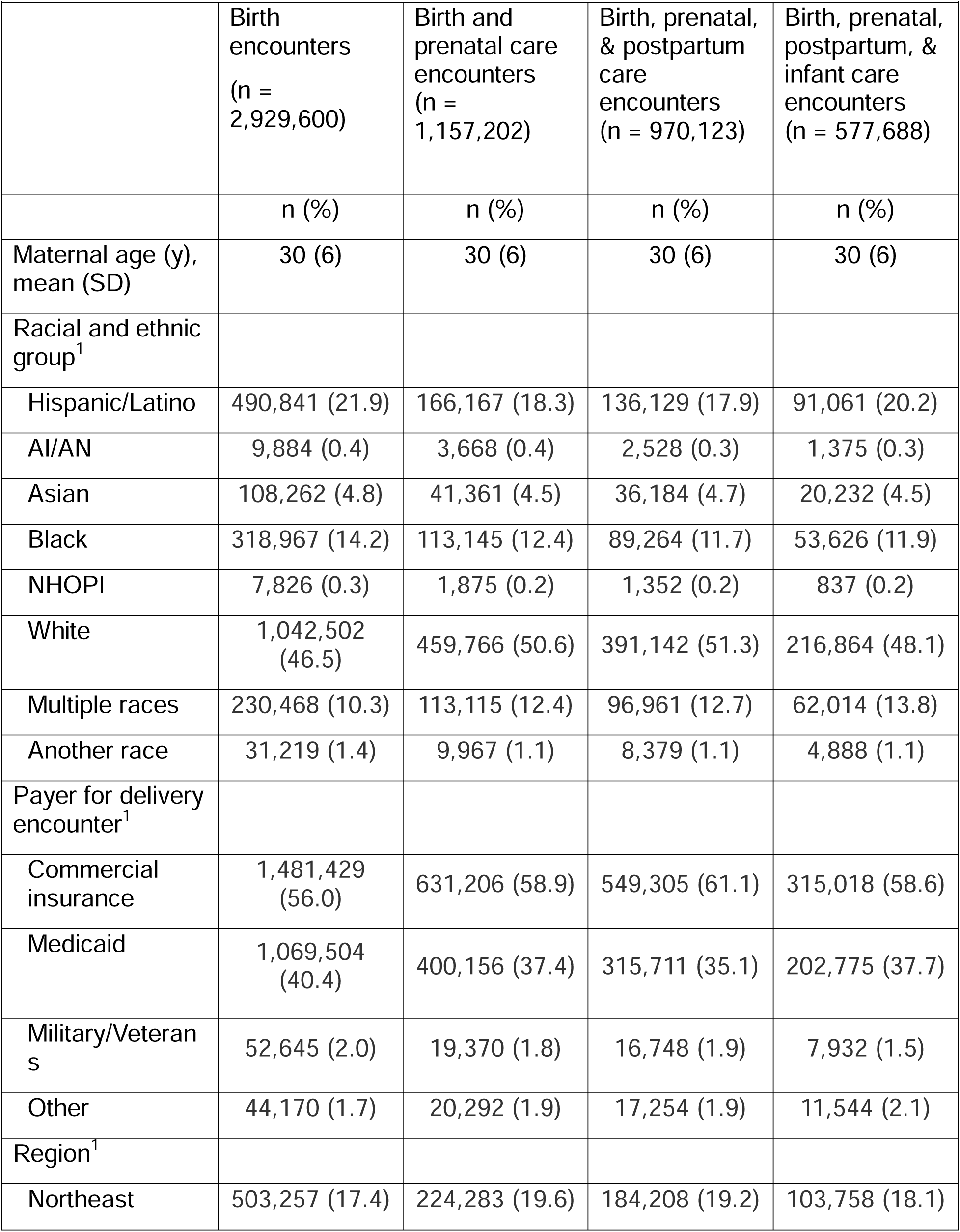
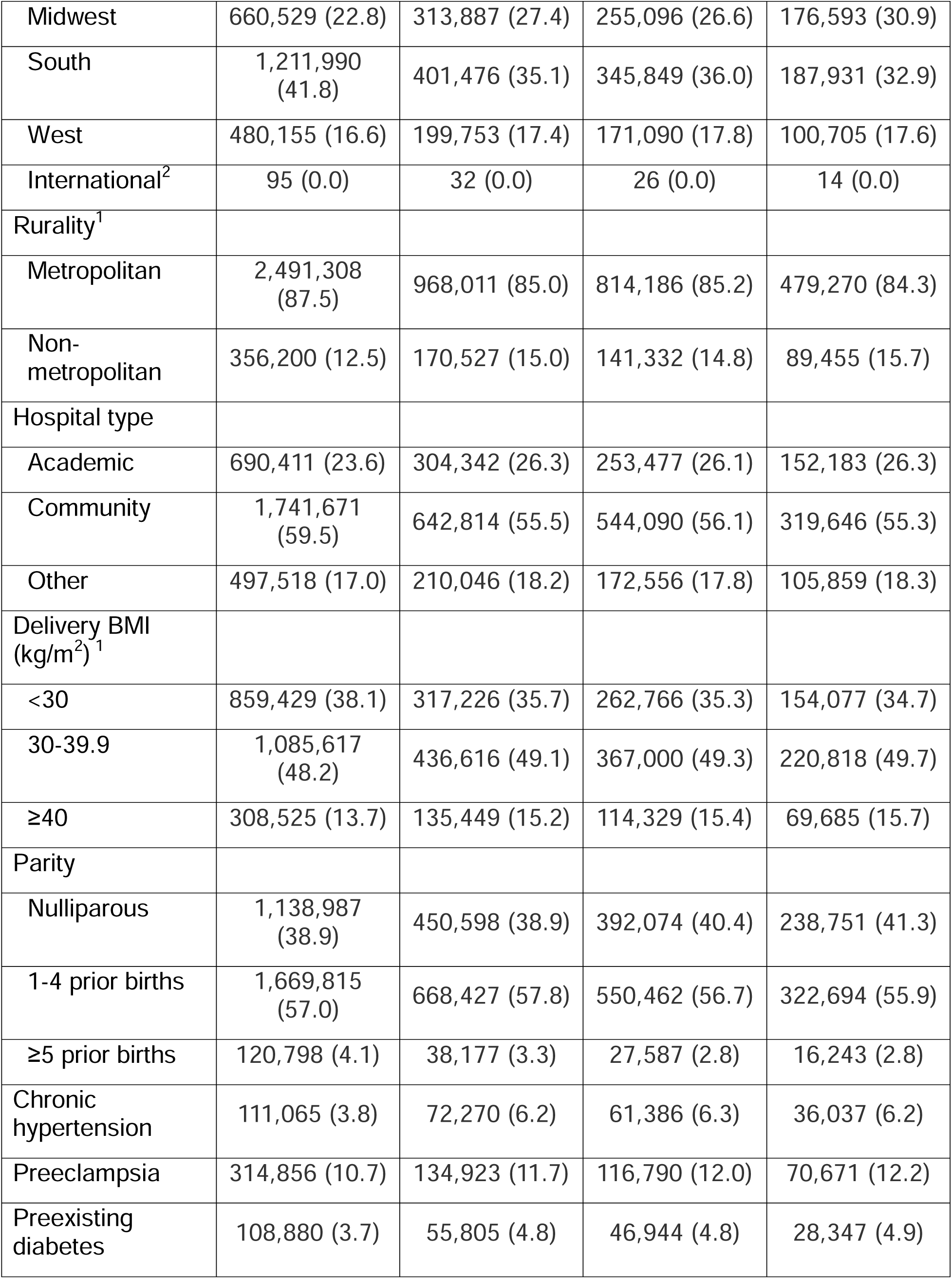
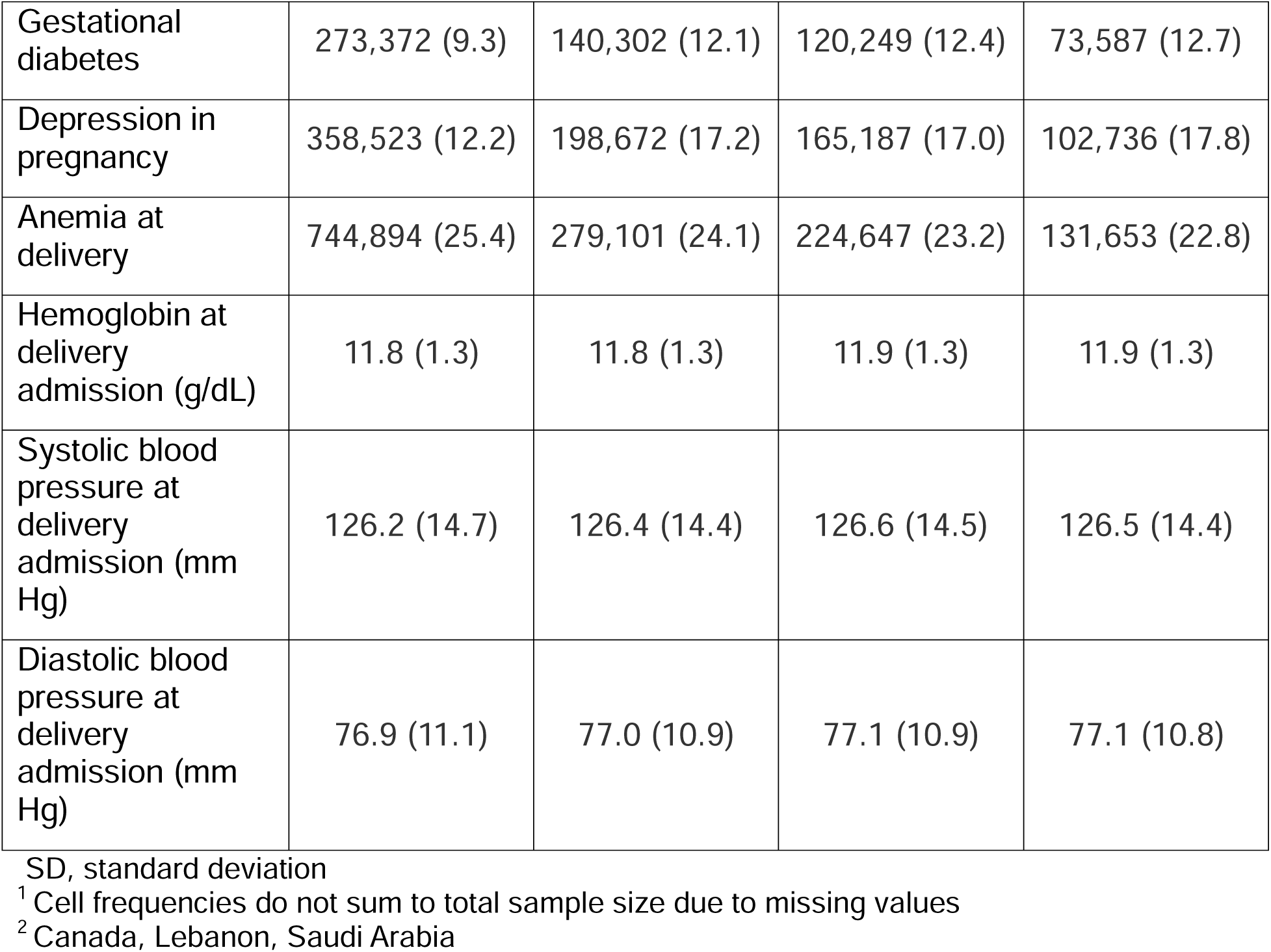
Maternal health characteristics of pregnancies by linked maternal-infant cohort selection

Birth and infant characteristics were also similar across the four cohorts (**Table 3**). An exception was lower prevalence of extremely preterm birth (<28 wk), very low birthweight (<1500 g), and very low 5-minute Apgar score (0-3) as well as very few in-hospital deaths in the cohort with an infant follow-up visit within 12 weeks of discharge from the birth hospitalization. In the total linked birth cohort, 3% of infants were twins, 33% were delivered by cesarean, 9% were born preterm (<37 wk), 9% had a low birthweight (<2500 g), and 2% had a 5-minute Apgar score <7.

**Table 3.**
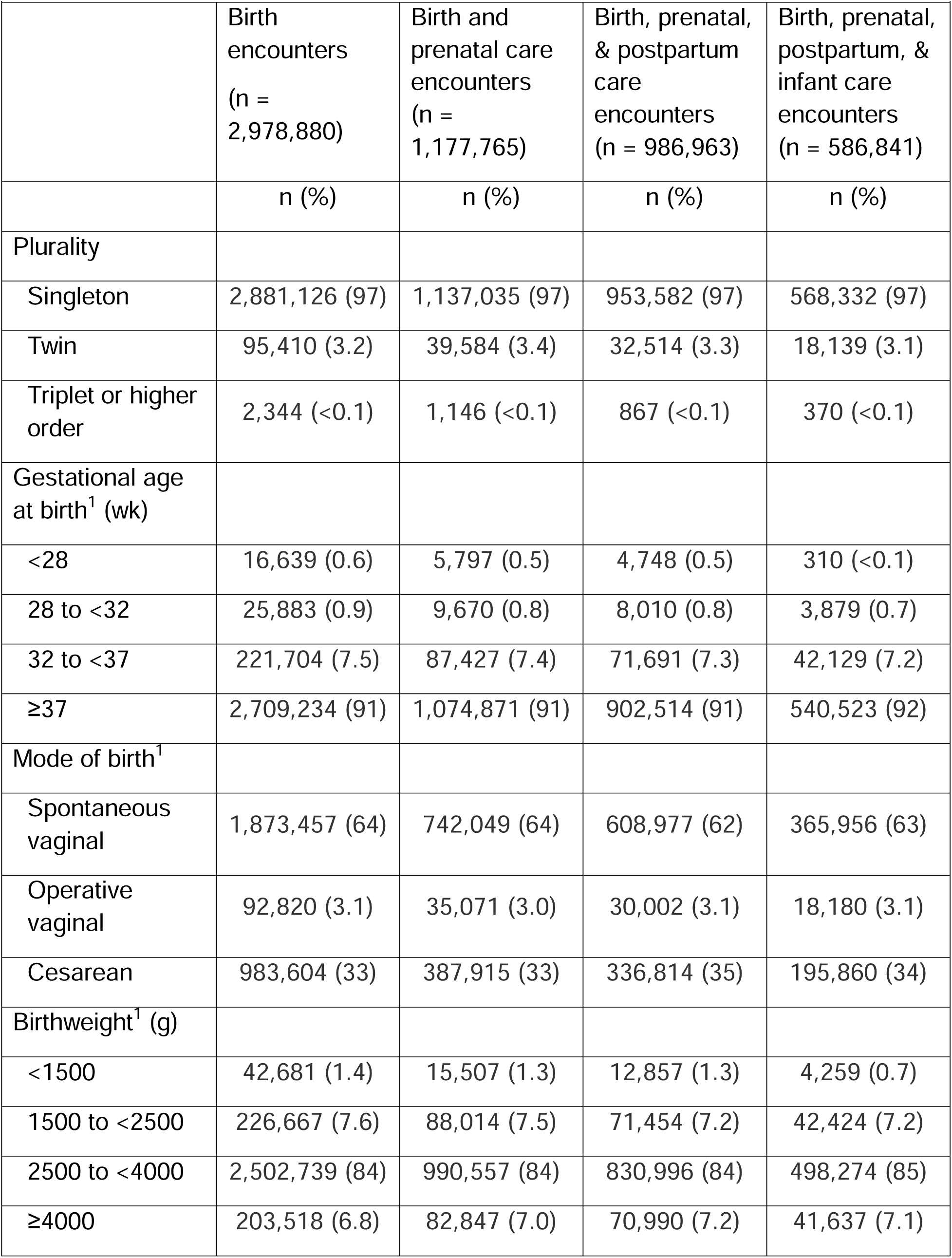
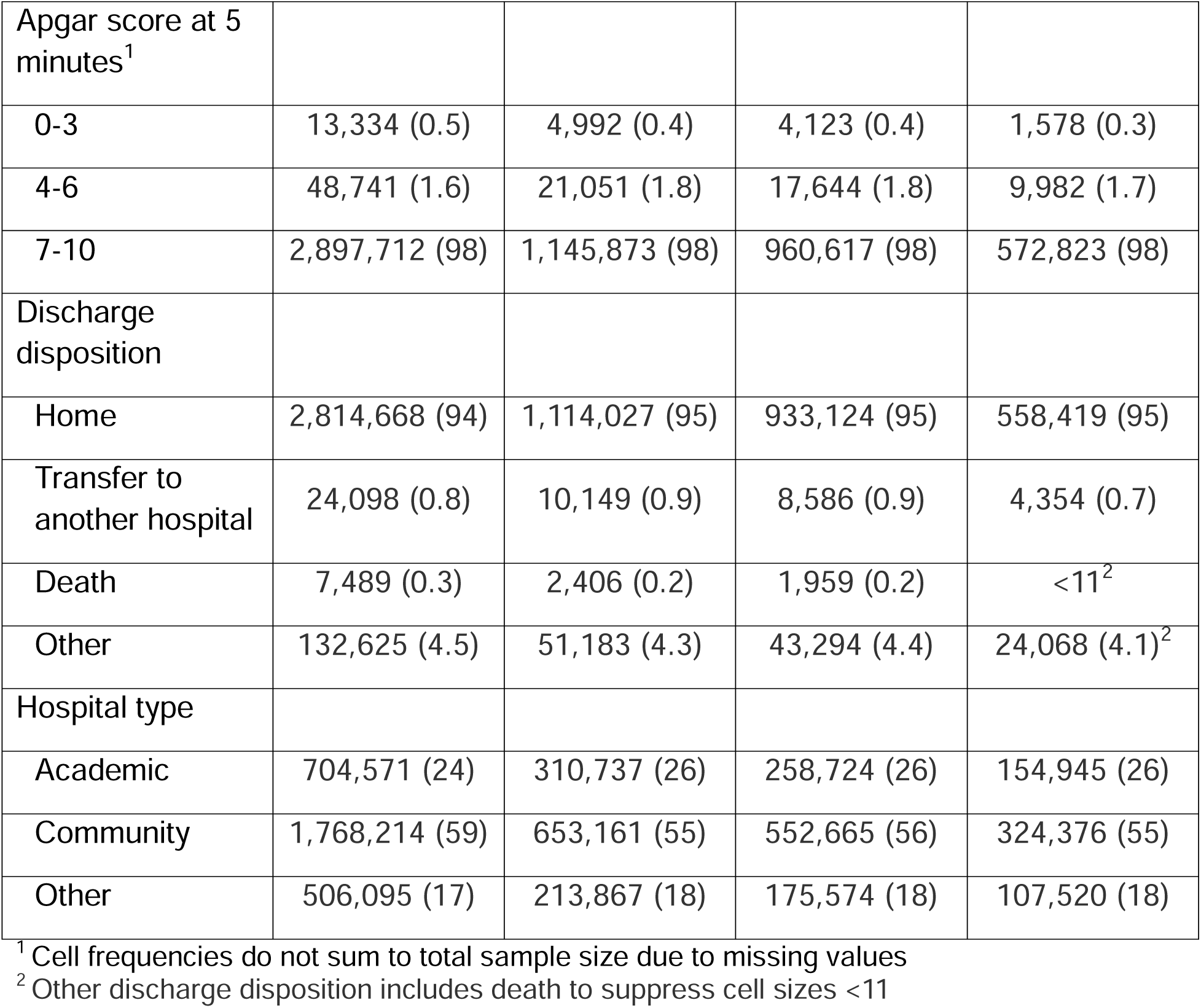
Characteristics of infants by linked maternal-infant cohort selection

|  | Birth encounters<br>(n = 2,978,880) | Birth and prenatal care encounters<br>(n = 1,177,765) | Birth, prenatal, & postpartum care encounters<br>(n = 986,963) | Birth, prenatal, postpartum, & infant care encounters<br>(n = 586,841) |
| --- | --- | --- | --- | --- |
|  | n (%) | n (%) | n (%) | n (%) |
| Plurality |  |  |  |  |
| Singleton | 2,881,126 (97) | 1,137,035 (97) | 953,582 (97) | 568,332 (97) |
| Twin | 95,410 (3.2) | 39,584 (3.4) | 32,514 (3.3) | 18,139 (3.1) |
| Triplet or higher order | 2,344 (<0.1) | 1,146 (<0.1) | 867 (<0.1) | 370 (<0.1) |
| Gestational age at birth<sup>1</sup> (wk) |  |  |  |  |
| <28 | 16,639 (0.6) | 5,797 (0.5) | 4,748 (0.5) | 310 (<0.1) |
| 28 to <32 | 25,883 (0.9) | 9,670 (0.8) | 8,010 (0.8) | 3,879 (0.7) |
| 32 to <37 | 221,704 (7.5) | 87,427 (7.4) | 71,691 (7.3) | 42,129 (7.2) |
| ≥37 | 2,709,234 (91) | 1,074,871 (91) | 902,514 (91) | 540,523 (92) |
| Mode of birth<sup>1</sup> |  |  |  |  |
| Spontaneous vaginal | 1,873,457 (64) | 742,049 (64) | 608,977 (62) | 365,956 (63) |
| Operative vaginal | 92,820 (3.1) | 35,071 (3.0) | 30,002 (3.1) | 18,180 (3.1) |
| Cesarean | 983,604 (33) | 387,915 (33) | 336,814 (35) | 195,860 (34) |
| Birthweight<sup>1</sup> (g) |  |  |  |  |
| <1500 | 42,681 (1.4) | 15,507 (1.3) | 12,857 (1.3) | 4,259 (0.7) |
| 1500 to <2500 | 226,667 (7.6) | 88,014 (7.5) | 71,454 (7.2) | 42,424 (7.2) |
| 2500 to <4000 | 2,502,739 (84) | 990,557 (84) | 830,996 (84) | 498,274 (85) |
| ≥4000 | 203,518 (6.8) | 82,847 (7.0) | 70,990 (7.2) | 41,637 (7.1) |
| Apgar score at 5 minutes<sup>1</sup> |  |  |  |  |
| 0-3 | 13,334 (0.5) | 4,992 (0.4) | 4,123 (0.4) | 1,578 (0.3) |
| 4-6 | 48,741 (1.6) | 21,051 (1.8) | 17,644 (1.8) | 9,982 (1.7) |
| 7-10 | 2,897,712 (98) | 1,145,873 (98) | 960,617 (98) | 572,823 (98) |
| Discharge disposition |  |  |  |  |
| Home | 2,814,668 (94) | 1,114,027 (95) | 933,124 (95) | 558,419 (95) |
| Transfer to another hospital | 24,098 (0.8) | 10,149 (0.9) | 8,586 (0.9) | 4,354 (0.7) |
| Death | 7,489 (0.3) | 2,406 (0.2) | 1,959 (0.2) | <11<sup>2</sup> |
| Other | 132,625 (4.5) | 51,183 (4.3) | 43,294 (4.4) | 24,068 (4.1)<sup>2</sup> |
| Hospital type |  |  |  |  |
| Academic | 704,571 (24) | 310,737 (26) | 258,724 (26) | 154,945 (26) |
| Community | 1,768,214 (59) | 653,161 (55) | 552,665 (56) | 324,376 (55) |
| Other | 506,095 (17) | 213,867 (18) | 175,574 (18) | 107,520 (18) |

<sup>1</sup> Cell frequencies do not sum to total sample size due to missing values
<sup>2</sup> Other discharge disposition includes death to suppress cell sizes <11

The analysis of the association between high SBP in the first trimester and preeclampsia included 64,894 pregnancies. Of 2,929,600 pregnancies in the full cohort, 1,275,421 (44%) had a prenatal care encounter in the first trimester. Among the 78,201 women with a chronic hypertension diagnosis, 64,894 (83%) had SBP recorded at a first-trimester prenatal encounter. The prevalence of high SBP (≥140 mm Hg) was 28.4%. Among those with a high SBP, the prevalence of preeclampsia was 39.2%, compared with 30.3% among those without a high SBP (**Table 4**). The RR for the association between high SBP and preeclampsia was 1.29 (95% CI: 1.26, 1.33) in the unadjusted regression model and 1.26 (95% CI: 1.23, 1.30) in the adjusted regression model.

**Table 4.**
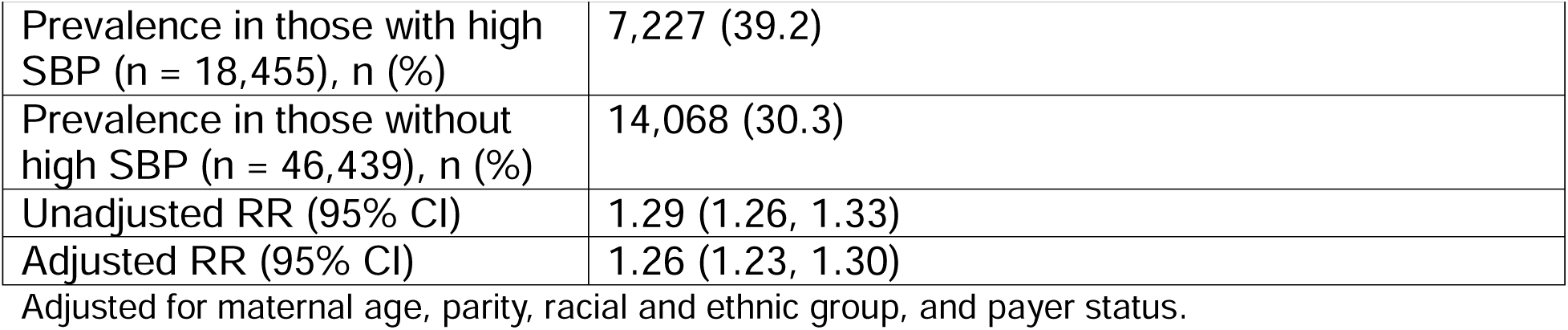
Association between high systolic blood pressure (SBP) (≥140 mm Hg) in the first trimester and the outcomes of preeclampsia among individuals with chronic hypertension (n = 64,894)

## COMMENT

This study demonstrated replicable steps for longitudinal, linked maternal-infant cohort creation using the Epic Cosmos dataset, found low missingness of key variables, and did not find evidence of selection bias with longitudinal cohort selection. In an applied analytical example using the cohort to replicate a known association, we found that high SBP (≥140 mm Hg) in the first trimester was associated with approximately 20-30% higher risk of developing preeclampsia among individuals with chronic hypertension. This example shows the utility of Cosmos for perinatal epidemiologic research questions that benefit from a very large, generalizable U.S.-based sample and utilize information recorded in structured EHR.

The Epic Cosmos maternal-infant cohort was found to be highly representative of the United States. Comparing the Cosmos 2023-2024 linked birth cohort to national birth certificate data from 2023,^16^ 42% vs 41% lived in the South, 23% vs 20% in the Midwest, 17% vs 16% in the Northeast, and 17% vs 21% in the West. Racial-ethnic composition was also comparable in the Cosmos and birth certificate cohorts, with the largest groups being 47% vs 50% non-Hispanic White, 22% vs 26% Hispanic/Latino, and 14% vs 14% non-Hispanic Black.^16^ The prevalences of common conditions during pregnancy, including gestational diabetes, anemia, and depression, were similar to those reported in other large studies in the United States.^17–20^ Diagnoses in EHR data may be prone to overreporting, such as coding for the purpose of ordering a diagnostic test, and improved phenotyping of perinatal conditions by Cosmos users for accurate identification of conditions and their onset is an important area of work that will be widely beneficial.^2^ We also encourage researchers to apply filters that restrict time windows for the diagnosis during pregnancy and differentiate mutually exclusive conditions (e.g., preexisting diabetes and gestational diabetes.)

Data completeness in the Epic Cosmos maternal-infant cohort was found to be high overall. Missingness was less than 1% for key perinatal characteristics often derived from birth certificates and unavailable in claims datasets (e.g., parity, birthweight, Apgar score, rurality). Missingness was higher (∼10%) for vital signs and laboratory results routinely measured at delivery admission (blood pressure and hemoglobin), and highest (∼24%) for racial-ethnic group and BMI at delivery. As demonstrated in our example analysis, we would encourage researchers to use missing data techniques in their analyses with Cosmos to reduce the risk of bias.

Sequential restrictions for longitudinal cohort selection across the prenatal, perinatal, and neonatal periods reduced cohort size but did not indicate that selection bias was induced, which is reassuring. However, researchers should be aware that longitudinal cohort selection will exclude certain patient groups (e.g., patients with delayed or no prenatal care or those who switch healthcare organizations). Although characteristics were similar, social selection bias may still be present with longitudinal restrcitions. These restrictions reduced cohort size considerably—from approximately 2.9 million pregnancies in the linked maternal-infant cohort to fewer than 600,000 pregnancies with longitudinal care from the first trimester through infant follow-up care. This reduction was likely driven by individuals who had a delivery at a Cosmos-participating healthcare organization but received prenatal or infant follow-up care at a healthcare organization that does not participate in Cosmos. Further, the number of source data organizations in Cosmos increases over time as more and more organizations adopt Epic EHR systems. Researchers should be aware of this and take steps to mitigate induced bias, such as by restricting to source healthcare organizations with data across a full study period when conducting trend analyses.

We conducted an example analysis in the study sample, finding that high SBP (≥140 mm Hg) in the first trimester was associated with approximately 25-30% increased risk of preeclampsia in later pregnancy among individuals with chronic hypertension. This association has been studied previously in selective samples,^9,21^ but lack of blood pressure information has precluded assessment in large datasets. The prevalence of high SBP and superimposed preeclampsia observed in our study is consistent with other studies of pregnant women with chronic hypertension.^15,21^ There is substantial clinical interest in whether tighter blood pressure control in early pregnancy could mitigate preeclampsia onset and severity among individuals with chronic hypertension. Since the multi-site CHAP trial in 2022,^14^ obstetrics societies in the U.S. have lowered their recommended blood pressure for antihypertensive treatment from ≥160/110 mm Hg to ≥140/90 mm Hg.^22^ There is currently a debate as to whether the threshold for treatment should be lowered further to ≥130/80 mm Hg.^14^ Other studies have reported preeclampsia risk to increase with increasing blood pressure in early pregnancy, even among individuals without chronic hypertension.^10,23^

Additional findings from our study for researchers to note is that restriction to those with prenatal care and delivery encounters increased the prevalence of some comorbidities (e.g., antenatal depression from 12% to 17%). Further, restriction to those with infant follow-up care reduced the prevalence of extremely preterm birth, very low birthweight, and very low 5-minute Apgar score; these differences may reflect the exclusion of in-hospital neonatal deaths and transfer of high-risk newborns outside of Cosmos-participating organizations with this cohort restriction. In examining infant discharge disposition from the birth hospitalization, no differences were observed except for very few (<11) recorded deaths in the cohort with infant follow-up. We limited our study to births with over one year of follow-up data, which is an important consideration for studies examining outcomes of high-risk newborns, who will typically have long hospitalizations and may require hospital transfer.

This study has limitations. This initial study only included pregnancies that ended in a live birth and an area of future work is developing a robust approach to identify non-live births including stillbirths, spontaneous abortions and pregnancy terminations. This approach will be distinct and more complex as non-live births would not be captured in birth records and, based on our experience, may not be recorded in pregnancy records, depending on the types of any healthcare services received during pregnancy and the pregnancy loss. Another important consideration for our results is that we did not place restrictions on variables, such as removing outlier values or requiring multiple records to confirm a diagnosis. This was an intentional decision to make the cohort steps useful to a broad set of researchers, who would make such decisions in the analytical stage. For example, it would be possible in Cosmos for a researcher to require both a diagnosis of gestational diabetes and a positive 3-hour glucose tolerance test. Finally, it is important for researchers to know that Cosmos contains structured EHR data, including demographic information, vital signs, diagnoses, procedures, medications prescribed and administered, and laboratory test results. However, not all structured EHR data are currently available, including types of flowsheets and laboratory results that appear as a scanned document in a chart. Epic continues to map additional structured data into Cosmos and therefore available data domains increase over time. Unstructured EHR data, such as clinical notes, are also not currently available. The cohort was predominantly from the United States (98.5%) and therefore generalizability to other countries is unknown.

In this study, we presented a rigorous and reproducible approach to creating longitudinal, linked maternal-infant cohorts in Epic Cosmos—a relatively new EHR dataset with a very large sample size and granular information from structured EHR data. Our findings demonstrate overall high completeness of key variables, high representativeness of the United States, and characteristics that do not meaningfully change with longitudinal cohort selection. We demonstrated the utility of the cohorts by analyzing the association between a vital sign measurement (systolic blood pressure) and a clinical outcome (preeclampsia superimposed on chronic hypertension). Overall, the study suggests that Epic Cosmos is a highly valuable data resource for perinatal epidemiologic research, while also highlighting limitations of which researchers should be aware, including considerable attrition with longitudinal cohort restrictions.

## Data Availability

Access to the study dataset, Epic Cosmos, is available through participating healthcare organizations. Details are available at https://cosmos.epic.com/.

## ACKNOWLEDGMENTS

Data used in this study came from Epic Cosmos, a dataset created in collaboration with a community of health systems using Epic representing more than 300 million patient records from over 2,000 hospitals and 47,000 clinics as of April 2026. The community represents patients from all 50 states, D.C., Canada, Lebanon, and Saudi Arabia. Further description of health systems participating in Cosmos is provided at https://cosmos.epic.com/community/. The Epic Cosmos team served as a knowledge resource in developing the methods presented in this article. Epic had no other role in the research conducted or the writing of the article.

## ETHICS APPROVAL STATEMENT

This study was determined to not be human subjects research by the Stanford Institutional Review Board.

