## Supplemental Tables for "Development of Longitudinal, Linked Maternal-Infant Cohorts using the Epic Cosmos Electronic Health Record Dataset"

**Appendix 1. Steps for creating the birth cohort.**

- Total birth cohort (CY 2023-2024)
  - Using infant birth records (generated for liveborn infant)
- Link to pregnancies
  - Join to pregnancy records (based on maternal records)
- Remove historical pregnancies
- Link to birth encounters during same birth years
- Remove unavailable birth encounters (i.e., missing, masked, deleted)
- Identify twins/multiples & assign first infant birth date as the pregnancy delivery date if multiple dates
  - Set the first infant birth date as the delivery date for the shared pregnancy (multiples infant birth dates may differ)
- Exclude any stillbirth pregnancies remaining in error (code present from pregnancy delivery date plus/minus 7 days)
  - ICD-10: O36.4XXO, O36.4XX1, O36.4XX2, O36.4XX3, O36.4XX4, O36.4XX5, O36.4XX9, Z37.1, Z37.3, Z37.4, Z37.6, Z37.7, P95
  - ICD-9: 656.40, 656.41, 656.43, V27.1, V27.3, V27.4, V27.6, V27.7, 768.1
- Assign gestational age (GA)
  - If multiple GAs for a pregnancy in the infant birth record, select lowest
  - If no GA, calculate as delivery date the estimated start date in the pregnancy record
    - The rest are set as missing
- Assign last menstrual period (LMP) (LMP = Delivery date – GA)

**Appendix 2. Details on variables created.**

Used ICD-10 and ICD-9 diagnosis codes in all diagnosis types (billing, encounter, admitting, discharge diagnosis or problem list) to identify disorders. Codes below indicate “starts with” (e.g., 648.0 includes 648.00-648.04) for brevity.

1. **Maternal characteristics**
   a. Maternal age at delivery from birth record
   b. Racial and ethnic group: from maternal patient demographic record; if Hispanic/Latino, classify as such, then classify all without Hispanic/Latino based on racial group. If ethnicity is missing, or not Hispanic/Latino and missing race then classify as missing.
   c. Payer for delivery encounter: from insurance coverage record
   d. Geography: from maternal patient demographic record of state of residence and categorized as: (1) Northeast; (2) Midwest; (3) South; (4) West; (5) Not U.S. Regions based on US Census Bureau statistical regions (<https://www2.census.gov/geo/pdfs/maps-data/maps/reference/us_regdiv.pdf> )
   e. Rurality: using USDA Rural-Urban Commuting Area Codes and classifications assigned by Epic
      i. Categorized as metropolitan (1-3), nonmetropolitan (4-10)
   f. Body mass index at delivery: from vital signs record
      i. Time window: [delivery date -7, delivery date]
      ii. When multiple exact time -> select earliest. If multiple on the soonest exact time choose the min value
      iii. Categorize for descriptive purposes as <30 kg/m^2^; 30 to <40 kg/m^2^; ≥40 kg/m^2^
   g. Parity (prior live births)
      i. (1) nulliparous; (2) 1-4 prior births; (3) 5+ prior births
   h. Chronic hypertension [LMP-90 d, LMP+140 d]
      i. ICD10: I10, O10.0, O11
      ii. ICD9: 401, 642.0, 642.7
   i. Preeclampsia [LMP+140 d, delivery date + 14 d]
      i. ICD10: O11, O14.0, O14.1, O14.2, O15
      ii. ICD9: 642.4, 642.5, 642.6, 642.7
   j. Preexisting diabetes mellitus [LMP, delivery date ]
      i. ICD10: O24.0, O24.1, O24.3, O24.8, Z79.4, E08-E13
      ii. ICD9: 250
   k. Gestational diabetes mellitus [LMP+210 days, delivery date]
      i. ICD10: O24.430, O24.434, O24.435, O24.439
      ii. ICD9: 648.80, 648.81, 648.82, 648.83, 648.84
      iii. Also required no code for preexisting diabetes
   l. Depression [LMP, delivery date ]
      i. ICD10: F32.0-F32.9, F32.A, F33.0-F33.9
      ii. ICD9: 296.2, 296.3, 311
   m. Anemia [delivery date -7 d, delivery date]
      i. Loinc codes: HCT 4544-3 or HGB 718-7
      ii. Hb <11 or Hct<33. Select first if multiple using the exact date if still multiple values on the same time, choose the min value.
   n. Hemoglobin measurement (mean, SD)
      i. [delivery date -7, delivery date ] - select first if multiple using the exact date if still multiple values on the same time, choose the min value.
   o. Blood pressure measurement during 1^st^ trimester prenatal encounter [LMP, LMP+97 d]
      i. Require during prenatal care encounter as defined elsewhere and select first if multiple using the exact date. If still multiple values, choose the min value.
   p. Blood pressure measurements at delivery admission

[delivery date -7, delivery date ]

i. Select first if multiple using the exact date. If still multiple values,

choose the min value.

2. **Pregnancy & infant characteristics**
   a. Plurality: (1) singleton; (2) twin; (3) triplet or higher order
   b. Gestational age at delivery: (1) <28 wk; (2) 28 to <32 wk; (3) 32 to <37 wk; (4) 37+ wk
   c. Mode of delivery: (1) spontaneous vaginal; (2) operative vaginal (forceps or vacuum); (3) cesarean
   d. Birthweight: (1) >=4000g; (2) 2500 to <4000 g; (3) 1500 to <2500 g; (4) <1500g
   e. Apgar score at 5 minutes: (1) 0-3; (2) 4-6; (3) 7-10

**Appendix 3. Steps for selection of longitudinal, linked maternal-infant cohorts.**

1. Starting with birth cohort as described in Appendix 1, additionally require ≥1 prenatal care encounter in 1^st^ trimester and ≥1 prenatal care in 2^nd^ trimester
   a. Prenatal care:
      - Encounter type is specified as prenatal visit or office visit with obstetrics and gynecology, midwifery, MFM, perinatology, family medicine, or primary care
   b. Time windows:
      - 1^st^ trimester: [LMP, LMP+97d]
      - 2^nd^ trimester: [LMP+98, LMP+195d]
2. Additionally require ≥1 postpartum care encounter
   a. Postpartum care:
      - Encounter type is specified as postpartum visit or office visit with obstetrics and gynecology, midwifery, MFM, perinatology, family medicine, or primary care
      - Time window: >delivery date, <delivery date+84d
3. Additionally require ≥1 infant follow-up encounter
a. Infant follow-up:
   - A healthcare encounter not at a hospital (not ER or inpatient) and with pediatrics or family medicine
b. Time window:
   - >birth hospitalization discharge data, <84d
